# Fourth Wave of COVID-19 in India : Statistical Forecasting

**DOI:** 10.1101/2022.02.23.22271382

**Authors:** Sabara Parshad Rajeshbhai, Subhra Sankar Dhar, Shalabh

## Abstract

The spread of COVID-19 pandemic has wave nature. This article proposes a statistical methodology to study and forecast the future waves. The methodology is applied to COVID-19 data from India to statistically forecast the occurrence of fourth wave in India. In the course of this study, the data is fitted by the mixture of Gaussian distribution, and Bootstrap methodology is used to compute the confidence interval of the time point of peak of the fourth wave. This methodology can also be used to forecast the fourth and other waves in other countries also.

## 1. Introduction

The first known case due to COVID-19 was first identified in December, 2019, and since then, the human race is suffering from the pandemic caused by the virus and its mutations. Many countries have already seen the third wave of the spread of this virus (e.g., India, Germany, USA and many more), and a few countries (e.g., South Africa and Zimbabwe) have started to face the fourth and higher waves of the COVID-19 pandemic. An important question that arises before the the scientists is how to forecast the possible occurrence of the COVID-19 wave, so that it can be found e.g., when the fourth wave is likely to happen in those countries yet to face the fourth wave. The third wave of COVID-19 was predicted for India using the concept of mixture of Gaussian distribution based on the data of Zimbabwe, see [5], and when the third wave in India is finishing, it is now clear that the forecast in [5] was almost correct, See Figure 1. Motivated by that study, we here investigate the forecasting of the fourth wave of COVID-19 outbreak in India.

**Figure 1:**
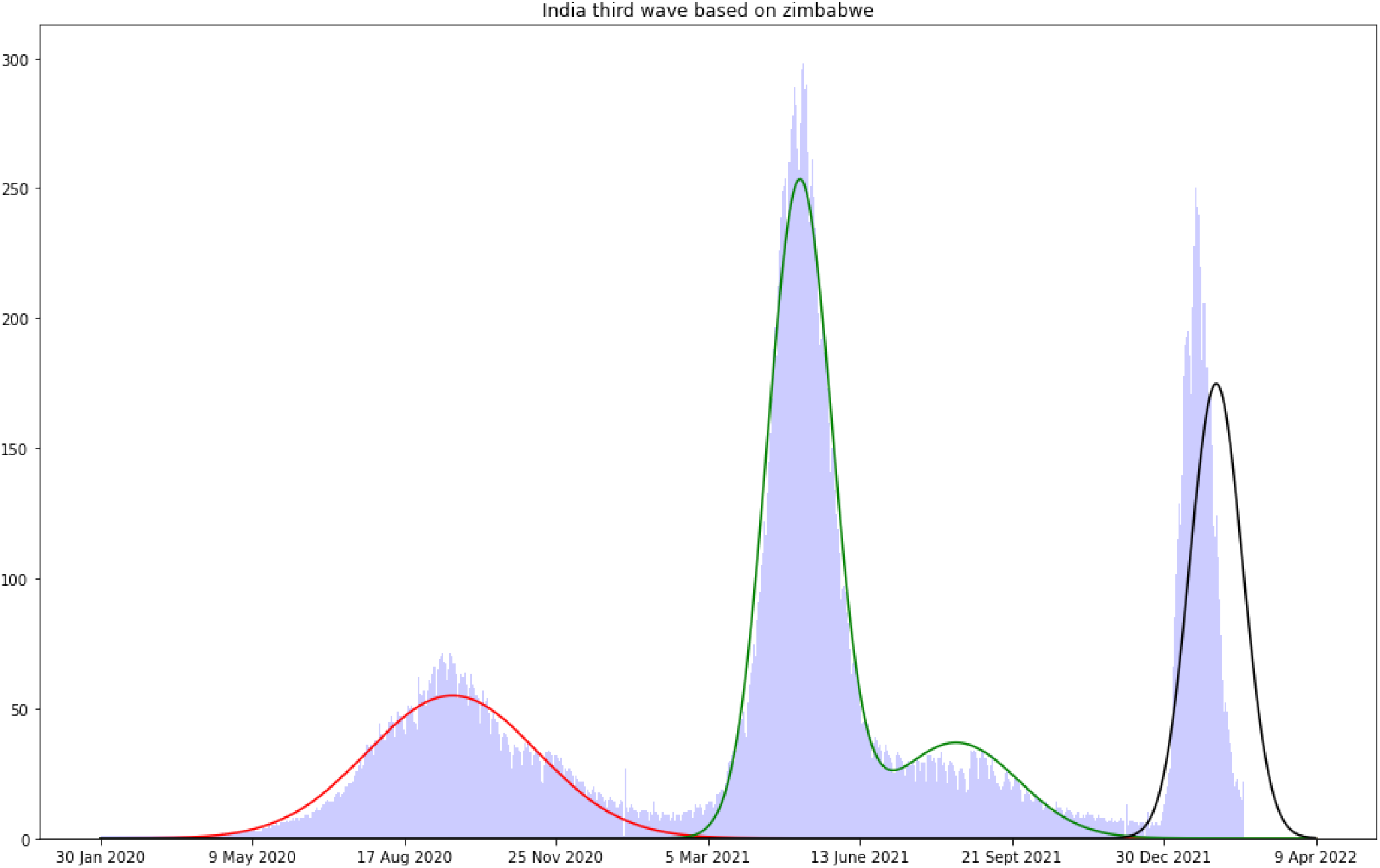
Prediction of India’s Fourth wave based on Zimbabwe

The researcher and scientists have modelled the COVID-19 waves using different approaches. Our approach is to use the data observed due to COVID-19 in the earlier waves in any country and train the model over that. Once the model is trained, the same is used to forecast the occurrence the next wave in other countries. The fundamental assumption in our study is that the behaviour of the SARS virus remains the same all over the world irrespective of countries. In other words, our thought process is that if the virus is causing the fourth wave in any country, why it will not cause the same in other country if the infection started spreading? As we mentioned in the preceding paragraph, there are a few countries, namely, South Africa and Zimbabwe already have started to face the fourth wave, and based on the data of those countries, one can predict the arrival and the end of the fourth wave in countries like India. We use the data on the number of COVID-19 cases of different COVID-19 waves and plotting it gives a fair idea about choosing a distribution which can possibly be fitted. Afterwards, using the concepts of mixture of Gaussian distribution for the infected cases/deaths due to COVID, we model it by the mixture of Gaussian distributions. Moreover, people are also interested to know when the peak of the curve will arrive. The tentative date of peak can be predicted as point estimate or confidence interval, i.e., the predicted value at a point or the prediction in intervals. We have used the well-known Bootstrap technique (see, e.g., [6]), to estimate the confidence interval of the time point of peak of the curve. This article addresses all these aforementioned issues.

We now want to discuss a few issues that may also affect the arrival of the fourth wave. For instance, there is always a fair chance that a new variant of this virus may have an intense impact on the whole analysis. The intensity of the impact will depend on the various factors like infectibility, fatality etc. For example, it has been observed that the Omicron variant is more infectious than the Delta variant but less fatal, and consequently, less hospitalization due to Omicron is reported, though the number of infected cases was more during the third wave time. Apart from this fact, the effect of vaccinations - first, second or booster dosage-may also play a significant role on the possibility of infection, degree of infection and various issues related to the fourth wave. The findings from this study are aimed to help and sensitize the people. For example, a few countries including the Government of India have started to provide Booster dose to a section of people, which may reduce the impact of the fourth wave in a long run. Overall, it should be mentioned that the forecast may be affected by all these reasons in the future.

The rest of the article is organized as follows. Section 2.1 describes the data set and its source, and the fitting of the mixture of Gaussian distribution to the data is explained in Section 2.2. The construction of confidence interval of the time of peak is explored in Section 2.3, and using all these technique, the final outcome of the forecast on the fourth wave is reported in Section 2.4. Section 3 contains the future work, and the article ends with a few relevant references.

## 2. Methodology and Analysis

### 2.1. Description of the Data set

The data set used for the analysis and prediction of COVID-19 waves has been taken from the public repository named Our World in Data (https://ourworldindata.org/coronavirus), see [3]. This data set is updated daily and includes the daily data about the daily new cases, confirmed total cases, daily new tests and total tests conducted for COVID-19, daily new deaths and total deaths attributed to COVID-19, excess mortality as a percentage difference from previous years, data about hospital occupancy such as number of COVID-19 patients in ICU, total number of COVID-19 patients in hospitals, and weekly new admissions of COVID-19 patients in hospitals, vaccination data such as the number of new daily vaccinations and total vaccinations so far (single, double and booster doses administered. It also includes positive rate of COVID-19 and real-time estimate of the effective reproduction rate (R) of COVID-19. Moreover, the data set consists of the observations as absolute values, per million of population of country as well as smoothed values. This data is originally sourced by John Hopkins University from governments, national and sub-national agencies across the world and is maintained by a team at its Center for Systems Science and Engineering (CSSE). It includes the data about the above mentioned parameters of COVID-19 for different countries since January 22, 2020. As it is common for many real data, this data set also contains missing values due to unavailability of data from the Governments. To overcome this issue, any missing data point is replaced with the average of the values on the previous and the next day.

Availability of Data and Materials: The original data sets analyzed during the current study are downloaded from the public repository of Our World in Data [3], and they can be accessed at https://home.iitk.ac.in/~shalab/covid-paper-data/Covid4wave/original_data_Covid4wave.csv. For the analysis presented in this paper, the actual sub-data set used is available at https://home.iitk.ac.in/~shalab/covid-paper-data/Covid4wave/filtered_data_Covid4wave.csv, and the python code used for the analysis is attached as a supplementary file “code.pdf”. The analysis was done using Jupyter Notebook, which can be run in any python 3 environment.

Figure 2(a) shows India’s daily new cases per million population, Figure 2(b) shows the daily new deaths from India attributed to COVID-19, Figure 2(c) shows daily new vaccination data for India, Figure 2(d) shows the comparison between daily new COVID-19 tests done in orange and daily new cases (positive) in blue, Figure 2(e) shows the comparison between the daily new cases per 3 million of population reported in India vs the Stringency Index. The stringency index is a composite measure based on nine response indicators including school closures, workplace closures, and travel bans, re-scaled to a value from 0 to 100 (100 = strictest).

**Figure 2:**
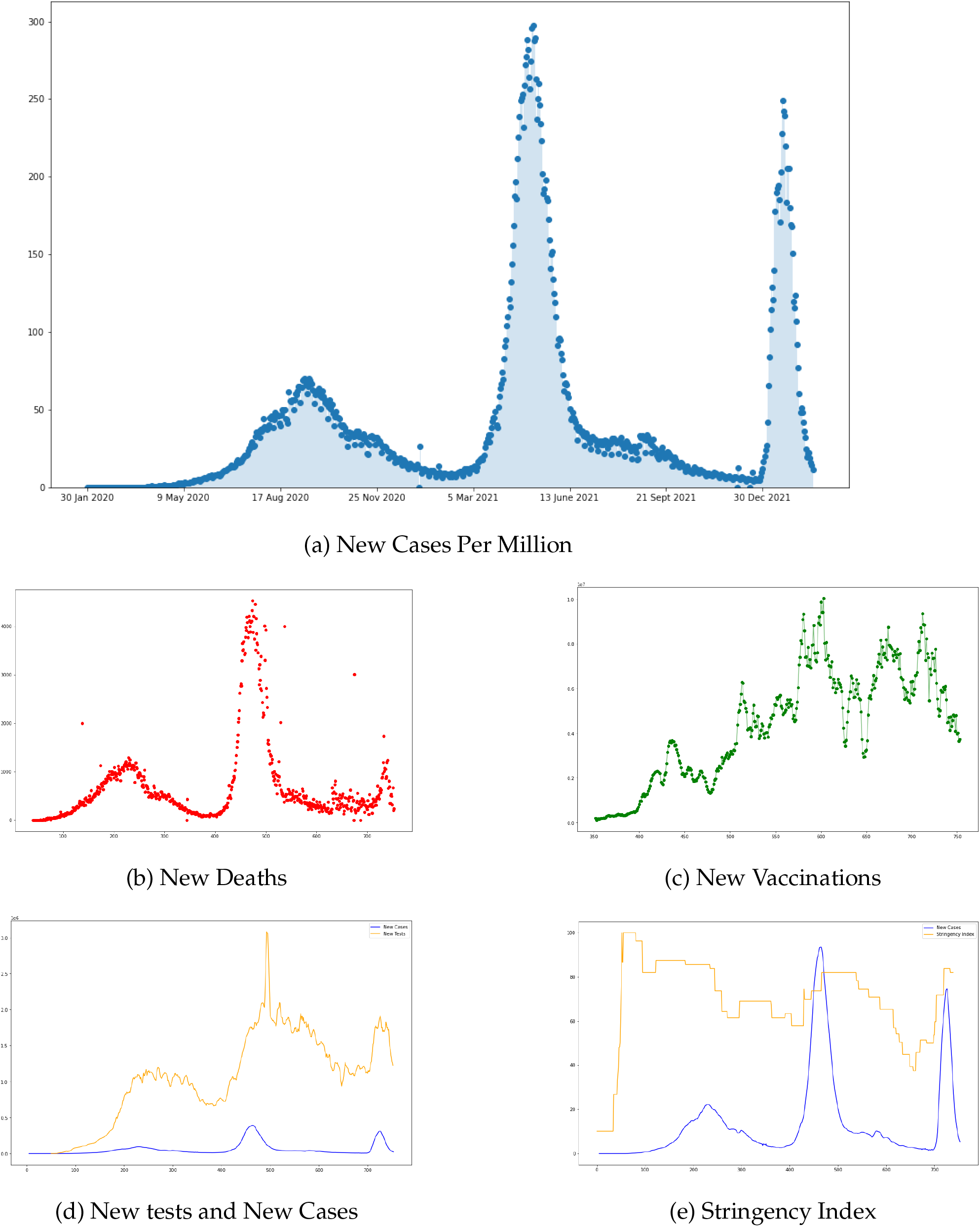
Visualizing the data set for India

### 2.2. Fitting Gaussian Mixture on the data

After checking and plotting the daily new cases of the first three waves of COVID-19 for India and other countries, it is clear that the different waves seem to be following some kind of multi-modal distribution. Hence, the conventional least squares methodology may not be able to achieve a good fit for the data. Also, the graphs for different countries seem to have some perturbations around the main waves. Therefore, a generalized approach involving polynomial regression model may not be possible due to the chance of over-fitting in some cases. However, the plots suggest that the individual waves seem to have similar plots, and hence, fitting a probabilistic model seems to be a good idea. Now, an important question is which probabilistic model will be a better fit to the data? Also, looking at the waves of India and other countries, the distribution of daily cases look like they are normally distributed around the peak dates of individual waves. Let us now define the Gaussian distribution.

For a real valued random variable *X* following a Gaussian Distribution, we can write: Suppose that *X* ∈ ℝ follows a Gaussian distribution with mean *μ* and standard deviation *σ*, i.e., symbolically writing, *X* ∼ 𝒩 (*μ, σ*). For *x* ∈ ℝ,

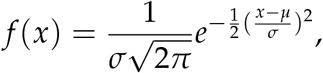

where *f* (.) is the probability density function.

If we consider a combination of first three waves of India, it then can be modelled as a mixture of three Gaussian distributions. Now, assuming that the daily cases data for India follows a mixture of three Gaussian Distribution, and suppose that the mean of the first wave, second wave and the third wave are *μ*_1_, *μ*_2_ and *μ*_3_, respectively, the standard deviation of the three waves are *σ*_1_, *σ*_2_ and *σ*_3_ respectively, and the individual weights of each waves are *w*_1_, *w*_2_ and *w*_3_, respectively. Hence, a mixture of Gaussian distributions can be a good model to fit on the daily cases data of COVID-19. The Gaussian mixture model (GMM) is well-known as an unsupervised learning algorithm for clustering, see [4], and it assumes that the underlying data points come from multi-dimensional Gaussian distributions that could have different unknown parameters. Here the objective is to find the unknown parameters of the Gaussian distributions that best explain our data, which is known as generative modeling. Overall, we are assuming that these data for each wave are normally distributed, and we want to find parameters that maximize the likelihood of observing these data. In other words, considering each data as being generated by a mixture of Gaussian distributions, its probability can be calculated, see e.g., [1], [2]. In notation,

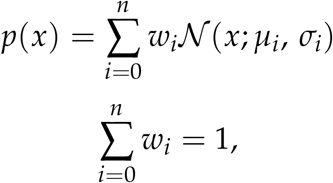

where *p*(*x*) is the probability density function, which is evaluated at *x*, of the random variable associated with number of daily new cases.

According to the first equation, the probability density function at any given point *x* will be a linear combination of *n* many certain normal distributions evaluated at *x* with their weight factors as *w*_*i*_, which denotes the strength of the *i*^*th*^ Gaussian distribution on the data point *x*, where *i* = 1, …, *n*. In view of the fact that *p*(*x*) is a probability density function, the individual weight factors *w*_*i*_ have a sum of 1. There are three different parameters that needs to be updated for each individual distribution: the means of the Gaussian distributions, i.e., *μ*_*i*_, the variances of each Gaussian, i.e., *σ*_*i*_, and the weights for each Gaussian, i.e., *w*_*i*_.

Note that in order to obtain the estimates of the unknown parameters, it turns out that we require the values of *w*_*i*_ to be known beforehand. This means that if we know the contribution factor of each individual distribution on a given data point, estimating the mean and the variance will be tractable. However, it is not true in this analysis as for any given date, the new case registered belongs to the first or the second or the third wave. There are two important points to take care of in the Gaussian mixture model in the course of study. First is to estimate the parameters for each Gaussian component within the Gaussian mixture, and the other one is to determine the contribution of the marginal Gaussian distributions to any data point. To perform it, Gaussian Mixture object of scikit-learn package of python [4] was used.

Gaussian Mixture models work based on an algorithm called Expectation-Maximization (EM) (see, e.g., [7]). Strictly speaking, given the number of clusters for a Gaussian Mixture model, the EM algorithm tries to figure out the parameters of these Gaussian distributions in two basic steps:

1. The E-step makes a guess of the parameters based on available data. Data points are assigned to a Gaussian cluster, and the probabilities are calculated that they belong to that cluster. The probability for a given Gaussian can be computed as: *w*_*i*_𝒩 (*x*; *μ*_*i*_, *σ*_*i*_), and for normalization, dividing by 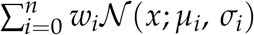
2. The M-step updates the cluster parameters based on the calculations from the E-step. The mean, variance and density are calculated for clusters based on the data points in the E-step. It is to be noted that the Gaussian Mixture Model model of sklearn in Python uses covariance as the call parameter, and it returns a matrix of variances of individual Gaussian distributions, see e.g., [2], [4]. To update the weight *w*_*i*_, one can sum up the probability that each point was generated by the *i*-th Gaussian distribution and divide by the total number of points. For the mean *μ*_*i*_, one can compute the mean of all points weighted by the probability of that point being generated by Gaussian *i*. For updating the variance Σ_*i*_, one can compute the variance of all points weighted by the probability of that point being generated by *i*-th Gaussian distribution. Finally, perform all these for each Gaussian distribution.

The two steps are repeated until convergence is reached.

### 2.3. Construction of Confidence Intervals

The confidence interval for the analysis is carried out using the Bootstrap method, which is a well-known re-sampling method. Precisely speaking, it consists of the re-sampling the original sample with replacement (Bootstrap Sample) and generating the Bootstrap replicates. At the first step, it generates bootstrap samples from the original sample by randomly choosing the observations among the original sample. Afterwards, it applies on the summary statistics such as variation, standard deviation, mean, and so forth to get replicates. We will use ‘mean’ to generate our bootstrap replicates. A major benefit of using Bootstrap re-sampling technique is that there is no need to assume the distributions of the observations. For this analysis, the 99% confidence interval is obtained by generating 15000 re-samples each having the same size as the original data.

### 2.4. Prediction

As we observed in Figure 3 that Zimbabwe and India have the maximum visible similarities in the shape of the COVID waves, the COVID cases in Zimbabwe is considered as the training data set. Further, assuming that the fourth wave of COVID-19 in India will follow a Gaussian Distribution, the parameters for the fourth wave come out to be:

**Figure 3:**
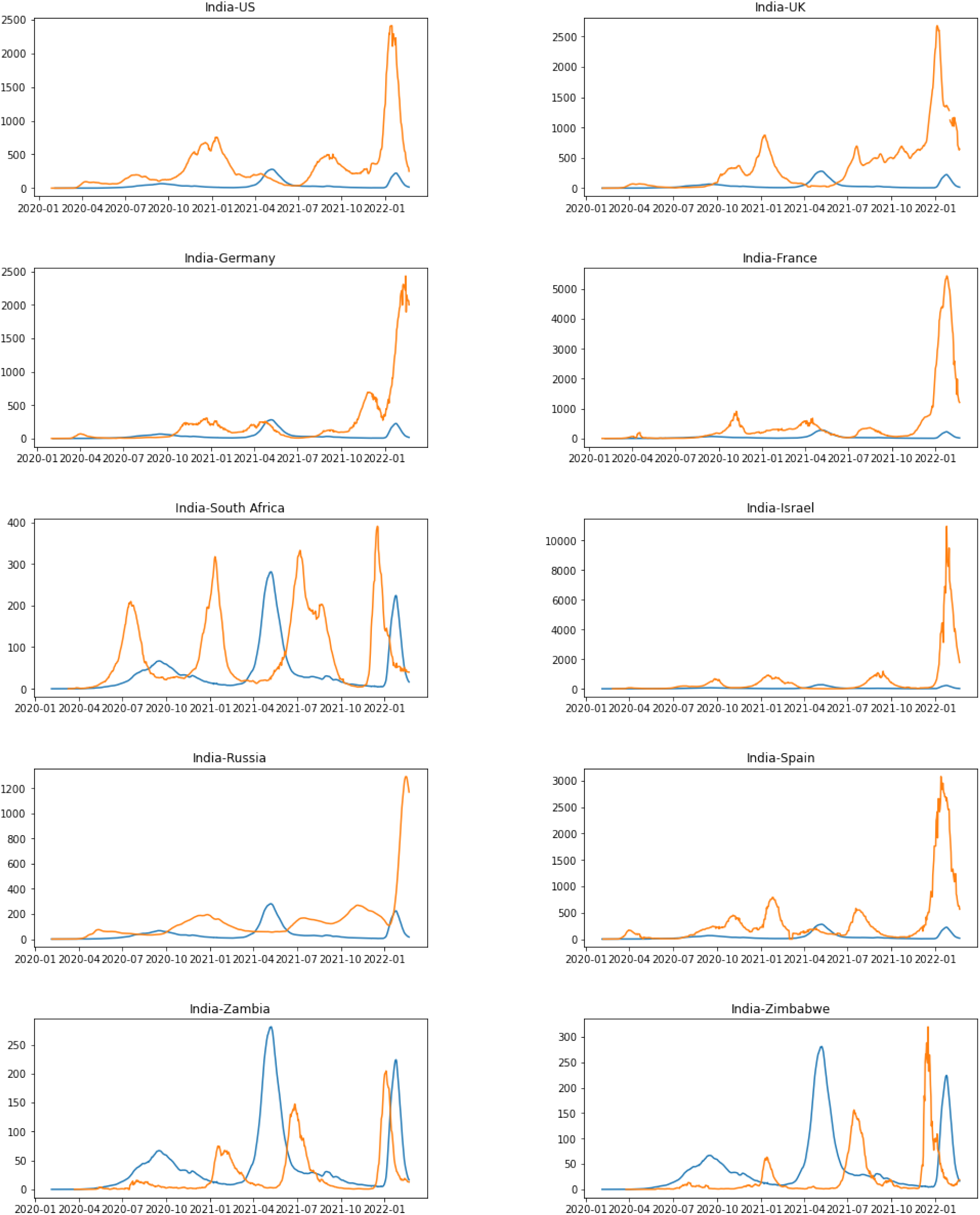
Prediction of India’s Fourth wave based on COVID data of Zimbabwe as the training data set.

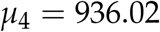

and

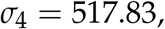

where (928.17, 943.86) is the 99% Bootstrap confidence interval for the mean *μ*_4_.

It indicates that the fourth wave of COVID-19 in India will arrive after 936 days from the initial data availability date, which is January 30, 2020. Therefore, the fourth wave starts from June 22, 2022, reaching its peak on August 23, 2022 and ends on October 24, 2022. Moreover, the 99% confidence interval for the date, when the curve will reach the peak, is approximately from August 15, 2022 to August 31, 2022. Figure 4 shows the prediction of the fourth wave of COVID-19 in India with fourth wave in black.

**Figure 4:**
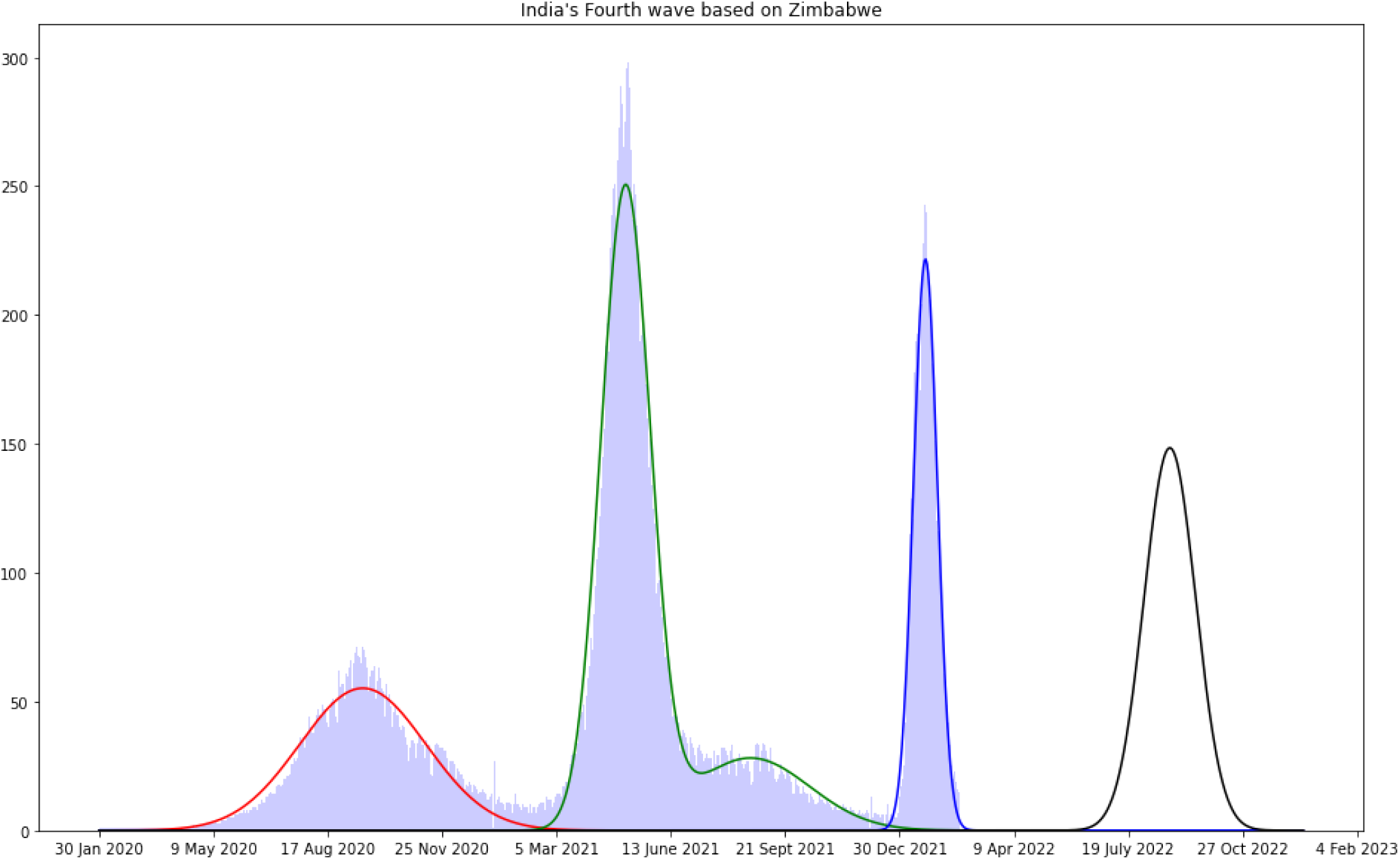
Prediction of India’s Fourth wave based on COVID data of Zimbabwe as the training data set.

## 3. Conclusion

### 3.1. Future Work

This article studies the forecasting of the fourth wave of COVID-19 outbreak in India. To investigate this research problem, the concept of fitting the mixture of Gaussian distribution to the data is used. Besides, the concept of the well-known Bootstrap technique is adopted to obtain the confidence interval of the time point of peak of the curve.

First of all, one may use other Statistical techniques like fitting the regression model to the data and predict the future possibilities using the regression model. Next, to construct the confidence interval, the methodology based on quantile regression can be adopted. As a future work, how the fatality of COVID virus is changing over time is a potential research problem.

### 3.2. Codes

The codes are available upon the request to the corresponding author.

## Data Availability

All data produced are available online at
https://home.iitk.ac.in/∼shalab/covid-paper-data/Covid4wave/original_ data_Covid4wave.csv
and
https://home.iitk.ac.in/∼shalab/covid-paper-data/Covid4wave/filtered_ data_Covid4wave.csv

https://ourworldindata.org/coronavirus

## ^1^Acknowledgement

The authors gratefully acknowledge the support from the MATRICS project from Science and Engineering Research Board (SERB), Department of Science and Technology, Government of India.

